# Waning of SARS-CoV-2 vaccine-induced immunity: A systematic review and secondary data analysis

**DOI:** 10.1101/2022.07.04.22277225

**Authors:** Francesco Menegale, Mattia Manica, Agnese Zardini, Giorgio Guzzetta, Valentina Marziano, Valeria d’Andrea, Filippo Trentini, Marco Ajelli, Piero Poletti, Stefano Merler

## Abstract

**Background:** The emergence of Omicron (B.1.1.529) variant of SARS-CoV-2 in late 2021 was followed by a marked increase of breakthrough infections. Estimates of vaccine effectiveness (VE) in the long term are key to assess potential resurgence of COVID-19 cases in the future.

**Methods:** We conducted a systematic review of manuscripts published until June 21, 2022 to identify studies reporting the level of protection provided by COVID-19 vaccines against SARS-CoV-2 infection and symptomatic disease at different time points since vaccine administration. An exponential model was used to perform a secondary data analysis of the retrieved data to estimate the progressive waning of VE associated with different vaccine products, numbers of received doses, and SARS-CoV-2 variants.

**Findings:** Our results show that VE of BNT162b2, mRNA-1273, ChAdOx1 nCoV-19 vaccines against any laboratory confirmed infection with Delta might have been lower than 70% at 9 months from second dose administration. We found a marked immune escape associated with Omicron infection and symptomatic disease, both after the administration of two and three doses. The half-life of protection against symptomatic infection provided by two doses was estimated in the range of 178-456 days for Delta, and between 66 and 73 days for Omicron. Booster doses were found to restore the VE to levels comparable to those acquired soon after administration of the second dose; however, a fast decline of booster VE against Omicron was observed, with less than 20% VE against infection and less than 25% VE against symptomatic disease at 9 months from the booster administration.

**Conclusions:** This study provides a cohesive picture of the waning of vaccine protection; obtained estimates can inform the identification of appropriate targets and timing for future COVID-19 vaccination programs.

## Introduction

The protection against SARS-CoV-2 infection and severe illness has progressively increased thorough the course of the pandemic, especially thanks to the massive vaccination programs that are being performed around the globe [1,2]. However, the replacement of the SARS-CoV-2 Delta variant by the Omicron variant, which took place around the globe in late 2021-early 2022, has been associated with a marked increase of breakthrough infections among vaccinated individuals [3-5], opening the debate on the effectiveness of vaccination against novel SARS-CoV-2 variants. As countries move into the third year of the COVID-19 pandemic, there is an urgent need to estimate the progressive waning of vaccine-induced protection associated with different vaccines and number of received doses. Several studies have quantified vaccine effectiveness (VE) against SARS-CoV-2 infection and symptomatic disease [6-21], but the obtained estimates are hard to reconcile.

In this study, we performed a systematic literature review of studies reporting VE at different time points since vaccine administration to estimate the waning of vaccine protection. We focused this review on the three most distributed COVID-19 vaccines in Western countries as of June 2022 [22]: BNT162b2 (Pfizer-BioNTech COVID-19 vaccine), mRNA-1273 (Moderna COVID-19 vaccine), and ChAdOx1 nCoV-19 (Oxford-AstraZeneca COVID-19 vaccine). We then performed a secondary analysis of the collected data to provide a cohesive picture of the waning rate associated with different vaccine products and quantified VE against SARS-CoV-2 infection and disease at any time from last dose administration, for different numbers of received doses, and different SARS-CoV-2 variants.

## Methods

### Search strategy and data selection

We searched PubMed and Web of Science for manuscripts providing evidence on the potential waning of COVID-19 VE over time. Predefined search terms were the following: (“Efficacy” OR “Effectiveness”) AND (“Vaccine” OR “Vaccination”) AND (“SARS-CoV-2” OR “COVID-19”) AND (“infection*” OR “disease”) AND (“waning” OR “decreas*”). Titles and abstracts of peer-reviewed articles and preprints published in English were screened from databases’ inception until June 21, 2022, to identify manuscripts including estimates of VE against SARS-CoV-2 infection or against symptomatic disease. After removing duplicates, we excluded studies not related to VE, providing results on antibody titer levels only, or estimating incidence rate, risk, or odds ratios based on vaccinated individuals only (i.e., comparing risks outcomes of individuals vaccinated at different times from vaccine administration or during different periods). After this preliminary screening, we scrutinized the full texts of the remaining manuscripts to assess their eligibility for our study according to the following criteria: (1) including data and estimates of VE against SARS-CoV-2 infection or against symptomatic disease after the primary vaccination course (consisting of two doses for the considered COVID-19 vaccines), or after the administration of a booster dose; (2) either comparing incidence, odds, or risk ratios of infection or disease between vaccinated and unvaccinated individuals, or analyzing vaccinated individuals only but considering the first two weeks after the first dose administration as a proxy for unvaccinated subjects; (3) estimating VE for at least two well-defined time intervals (e.g., “from 3 to 4 weeks from vaccine administration”); (4) providing information on which variants were circulating during the VE assessment; (5) providing VE estimates at population level or for all age-groups eligible for vaccination.

Average estimates of the VE at different times from the administration of the last dose were retrieved from the original studies to inform a simple statistical model to estimate the progressive waning of immunity. To minimize potential biases led by the initial ramp-up of vaccine-induced protection, we excluded from our analysis data points associated with VE measured during the first 14 days following the administration of the considered dose. Data points associated with less than 20 infections observed in the vaccinated group were excluded from the analysis.

### Data analysis

To estimate the vaccine-induced protection at any time from last dose administration, we modeled the VE as an exponential decay function of time:

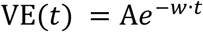

where *t* represents the number of days elapsed from the full activation of the vaccine protection (assumed to occur 14 days after the administration of any dose), *A* is the VE at 14 days after the administration of the last dose, *w* represents the waning rate associated with the vaccine-induced protection against the considered endpoint. Free model parameters (*A* and *w*) were estimated for each time series via a Markov chain Monte Carlo (MCMC) approach with Metropolis-within-Gibbs sampling algorithm applied to the normal likelihood of observing the original average values of VE estimated at different time intervals from vaccination. Model estimates associated with a specific time interval [*t*_1_, *t*_2_] were obtained by averaging the VE estimated over the considered period as 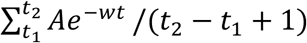. Once calibrated, the model was used to compare the estimated protection against different SARS-CoV-2 variants as provided by different vaccine products and number of administered doses at 14 days, and at 3, 6, and 9 months from last dose administration. Mean half-life of the vaccine-induced protection since last dose administration was computed as *log(2)/w+14* days. The considered modeling approach allowed us to compare the mean VE obtained from different studies at any time from last dose administration and to project VE in the longer term. Pooled estimates of the mean VE obtained from different studies were computed with the meta package using R statistical software version 4.1.2 (R Project for Statistical Computing).

## Results

We identified 544 original articles and 115 reviews published on peer-review journals, and 21 preprints (see Figure S1 in the Supplementary Material). Following screening of titles and abstracts, 39 studies reporting evidence of VE against SARS-CoV-2 infection and symptomatic disease were assessed for eligibility. We excluded 3 modeling studies, 8 manuscripts providing estimates for specific age-groups only, and 11 estimating the VE against unspecified SARS-CoV-2 variants. Finally, we excluded 5 studies providing an insufficient number of data points to perform our analysis (e.g., VE provided for <2 well-defined time intervals). Twelve articles [6-17] were included in our analysis. Original estimates of VE reported in these papers were obtained as result of test negative case-control studies [7,9,10,14-17], retrospective [8,12,13] or prospective cohort studies [6,11] assessing the difference in incidence of SARS-CoV-2 infection between vaccinated individuals and a certain reference group. Estimates extracted from [6-11,14-17] were obtained by using unvaccinated individuals as reference group. In [12] subjects vaccinated with one dose from less than 14 days were considered as a proxy for unvaccinated individuals. Similarly, in [13] the reference group was defined by subjects vaccinated with one dose from 4 to 10 days. According to the authors of [12,13], the rationale for this assumption was that unvaccinated people might undergo a higher number of tests and have their social habits altered by restrictions (e.g., EU Digital COVID certificate), therefore leading to biased VE estimates.

The articles included in this analysis provide estimates of VE over time of ChAdOx1 nCoV-19 (Oxford-AstraZeneca COVID-19 vaccine), BNT162b2 (Pfizer BioNTech COVID-19 vaccine), mRNA-1273 (Moderna COVID-19 vaccine) either against Alpha (B.1.1.7), Delta (B.1.617.2), or Omicron (B.1.1.529) variants [6-17]. Two studies [12,13] provide estimates of VE over time for unspecified mRNA vaccines, although BNT162b2 was prevalently adopted in the analyzed sample (85.2% and 66.2%, respectively). None of the analyzed studies found a temporal waning of VE of two doses against Alpha, nor of booster doses against Delta, possibly due to the shorter follow-up associated with the available records. From the selected papers, we extracted VE estimates associated with 1) two doses against any SARS-CoV-2 laboratory confirmed infection (asymptomatic or symptomatic) for Delta [6-13]; 2) two doses against symptomatic SARS-CoV-2 infection for Delta [15-17], and Omicron [16]; 3) two doses followed by a booster dose against any SARS-CoV-2 laboratory confirmed infection (asymptomatic or symptomatic) and against symptomatic infection for Omicron [14]. As a result, we considered 30 different time series, counting 155 data points associated with the VE of different vaccine products against different variants over time. The sample size, study period, type of study, variant, vaccine product, number of doses, and the endpoint associated with the analyzed time series of VE are summarized in Table S1 of the Supplementary Material.

The adopted modeling approach well captured the temporal changes in the mean values of VE reported by the original manuscripts (Figures 1 and 2), allowing a comparison of VE expected for different lineages, vaccine products, and number of administered doses over time and providing VE estimates in the long term (e.g., at 9 months from vaccination). Model estimates of the VE at 14 days from the vaccine administration (parameter *A*) and of the waning rate (*w*) associated with the different time series considered are reported in Table 1 and Table 2, along with the corresponding estimates obtained for the mean half-life of the vaccine-induced protection against the two considered endpoints.

**Table 1.** Model estimates of VE after the ramp-up (*A*), of the VE waning rate (*w*), and of the half-life of VE against any laboratory confirmed infection with Delta after the administration of two vaccine doses and with Omicron after the administration of a booster dose.

|  | VE at 14 days from last dose administration (%) [95% CI] | Waning rate (days<sup>-1</sup>) [95% CI] | Half life (days)* [95% CI] | Reference |
| --- | --- | --- | --- | --- |
| **2 doses against lab-confirmed infection for Delta** |  |  |  |  |
| **BNT162b2** | 91.7 [90.6 - 93.0] | 0.0009 [0.0008 - 0.0010] | 814.9 [721.6 - 910.1] | [6] |
|  | 93.7 [91.8 - 95.4] | 0.0008 [0.0006 - 0.0009] | 905.0 [795.7 - 1128.9] | [7]† |
|  | 92.3 [90.5 - 94.2] | 0.0008 [0.0006 - 0.0009] | 909.1 [751.5 - 1163.9] | [7]** |
|  | 94.3 [79.9 - 99.8] | 0.0037 [0.0009 - 0.0053] | 199.7 [144.0 - 786.0] | [8] |
|  | 94.7 [83.4 - 99.8] | 0.0101 [0.0078 - 0.0122] | 82.4 [70.8 - 102.6] | [9] |
| **mRNA-1273** | 94.8 [93.5 - 96.0] | 0.0009 [0.0007 - 0.0011] | 767.8 [635.3 - 959.6] | [6] |
|  | 97.9 [93.5 - 99.9] | 0.0016 [0.0013 - 0.0020] | 436.6 [362.6 - 556.9] | [7] † |
|  | 96.2 [88.6 - 99.9] | 0.0017 [0.0009 - 0.0025] | 415.4 [294.4 - 819.1] | [7]** |
|  | 96.3 [91.6 - 99.8] | 0.0014 [0.0009 - 0.0018] | 500.8 [393.0 - 754.1] | [10] |
|  | 90.0 [86.5 - 93.7] | 0.0010 [0.0006 - 0.0013] | 735.3 [566.6 - 1095.8] | [11] |
| **ChAdOx1 nCoV-19** | 81.6 [79.0 - 84.0] | 0.0006 [0.0003 - 0.0010] | 1085.8 [723.8 - 2443.4] | [6] |
|  | 78.0 [74.1 - 82.3] | 0.0007 [0.0002 - 0.0012] | 1022.8 [585.4 - 2945.0] | [7] † |
|  | 96.4 [88.1 - 99.9] | 0.0023 [0.0014 - 0.0028] | 317.0 [258.6 - 502.8] | [7]** |
| **Unspecified vaccine** | 91.6 [84.9 - 97.7] | 0.0050 [0.0042 - 0.0058] | 153.0 [134.4 - 180.4] | [12] |
|  | 87.8 [61.1 - 99.4] | 0.0066 [0.0036 - 0.0088] | 118.7 [92.7 - 204.7] | [13] |
| **Booster against lab-confirmed infection for Omicron** |  |  |  |  |
| **2 doses of BNT162b2 + BNT162b2** | 82.4 [71.5 - 91.6] | 0.0058 [0.0033 - 0.0081] | 132.5 [99.1 - 221.5] | [14] |

\*log(2)/ $w$ + 14 days
† Data from British Columbia
\*\* Data from Quebec

**Table 2.**
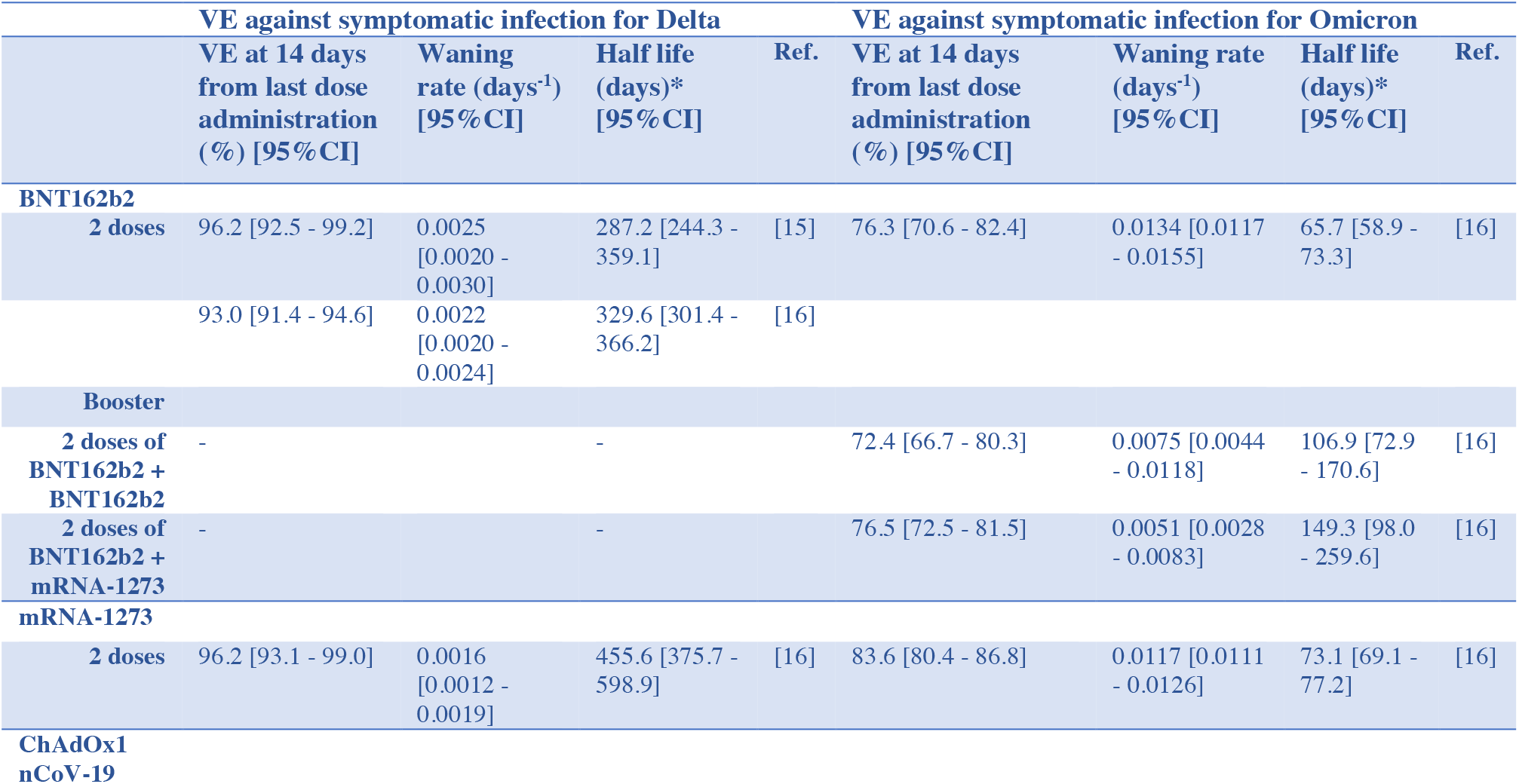
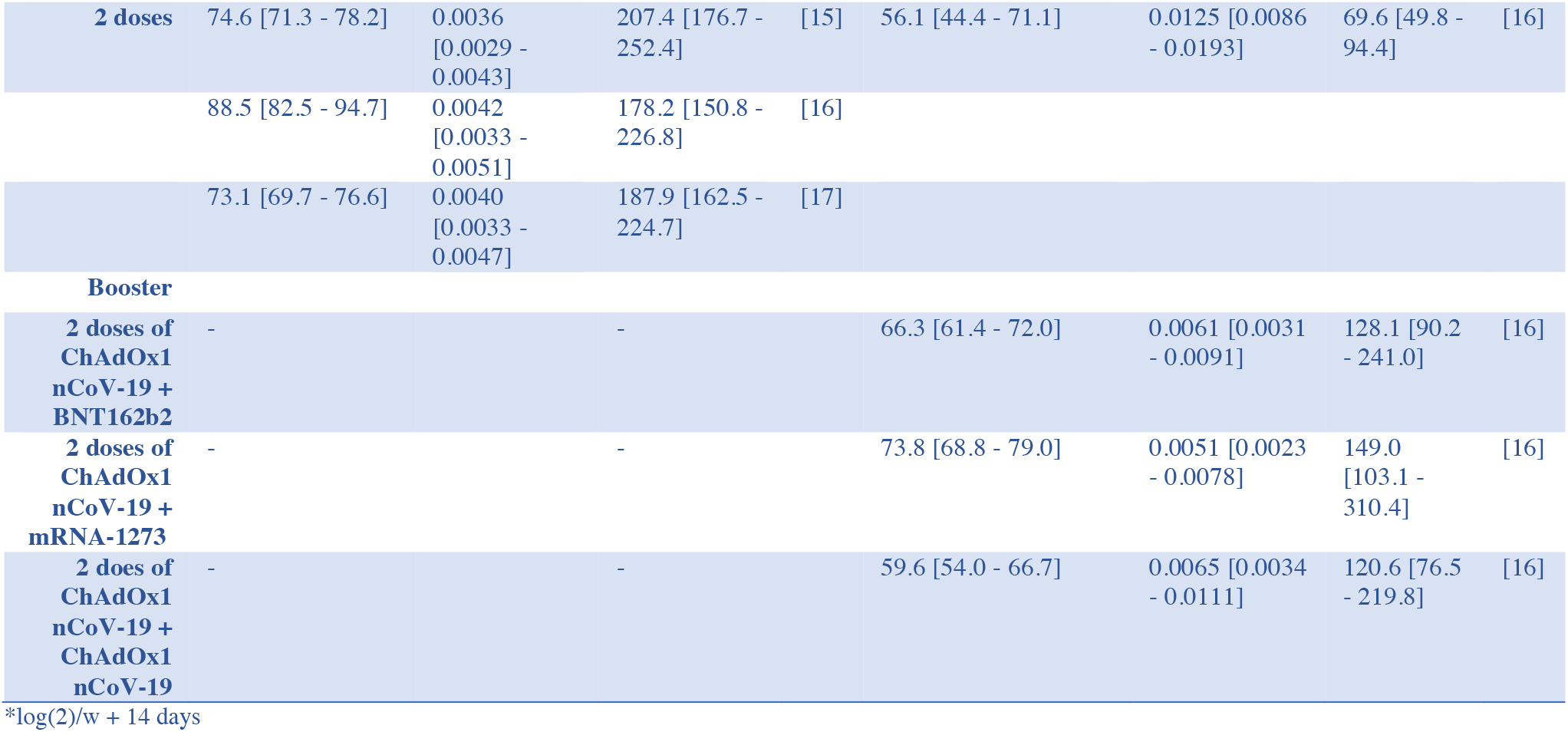
Model estimates of VE after the ramp-up (*A*), of the VE waning rate (*w*), and of the half-life of VE against symptomatic infection with Delta and Omicron after the administration of two vaccine doses and of a booster dose.

| VE against symptomatic infection for Delta |  |  |  |  | VE against symptomatic infection for Omicron |  |  |  |
| --- | --- | --- | --- | --- | --- | --- | --- | --- |
|  | VE at 14 days<br>from last dose<br>administration<br>(%) [95% CI] | Waning<br>rate (days<sup>-1</sup>)<br>[95% CI] | Half life<br>(days)*<br>[95% CI] | Ref. | VE at 14 days<br>from last dose<br>administration<br>(%) [95% CI] | Waning rate<br>(days<sup>-1</sup>)<br>[95% CI] | Half life<br>(days)*<br>[95% CI] | Ref. |
| BNT162b2 |  |  |  |  |  |  |  |  |
| 2 doses | 96.2 [92.5 - 99.2] | 0.0025<br>[0.0020 -<br>0.0030] | 287.2 [244.3 -<br>359.1] | [15] | 76.3 [70.6 - 82.4] | 0.0134 [0.0117<br>- 0.0155] | 65.7 [58.9 -<br>73.3] | [16] |
|  | 93.0 [91.4 - 94.6] | 0.0022<br>[0.0020 -<br>0.0024] | 329.6 [301.4 -<br>366.2] | [16] |  |  |  |  |
| Booster |  |  |  |  |  |  |  |  |
| 2 doses of<br>BNT162b2 +<br>BNT162b2 | - |  | - |  | 72.4 [66.7 - 80.3] | 0.0075 [0.0044<br>- 0.0118] | 106.9 [72.9<br>- 170.6] | [16] |
| 2 doses of<br>BNT162b2 +<br>mRNA-1273 | - |  | - |  | 76.5 [72.5 - 81.5] | 0.0051 [0.0028<br>- 0.0083] | 149.3 [98.0<br>- 259.6] | [16] |
| mRNA-1273 |  |  |  |  |  |  |  |  |
| 2 doses | 96.2 [93.1 - 99.0] | 0.0016<br>[0.0012 -<br>0.0019] | 455.6 [375.7 -<br>598.9] | [16] | 83.6 [80.4 - 86.8] | 0.0117 [0.0111<br>- 0.0126] | 73.1 [69.1 -<br>77.2] | [16] |
| ChAdOx1<br>nCoV-19 |  |  |  |  |  |  |  |  |
| **2 doses** | 74.6 [71.3 - 78.2] | 0.0036<br>[0.0029 - 0.0043] | 207.4 [176.7 - 252.4] | [15] | 56.1 [44.4 - 71.1] | 0.0125 [0.0086 - 0.0193] | 69.6 [49.8 - 94.4] | [16] |
|  | 88.5 [82.5 - 94.7] | 0.0042<br>[0.0033 - 0.0051] | 178.2 [150.8 - 226.8] | [16] |  |  |  |  |
|  | 73.1 [69.7 - 76.6] | 0.0040<br>[0.0033 - 0.0047] | 187.9 [162.5 - 224.7] | [17] |  |  |  |  |
| **Booster** |  |  |  |  |  |  |  |  |
| **2 doses of ChAdOx1 nCoV-19 + BNT162b2** | - |  | - |  | 66.3 [61.4 - 72.0] | 0.0061 [0.0031 - 0.0091] | 128.1 [90.2 - 241.0] | [16] |
| **2 doses of ChAdOx1 nCoV-19 + mRNA-1273** | - |  | - |  | 73.8 [68.8 - 79.0] | 0.0051 [0.0023 - 0.0078] | 149.0 [103.1 - 310.4] | [16] |
| **2 doses of ChAdOx1 nCoV-19 + ChAdOx1 nCoV-19** | - |  | - |  | 59.6 [54.0 - 66.7] | 0.0065 [0.0034 - 0.0111] | 120.6 [76.5 - 219.8] | [16] |

\*log(2)/w + 14 days

**Figure 1.**
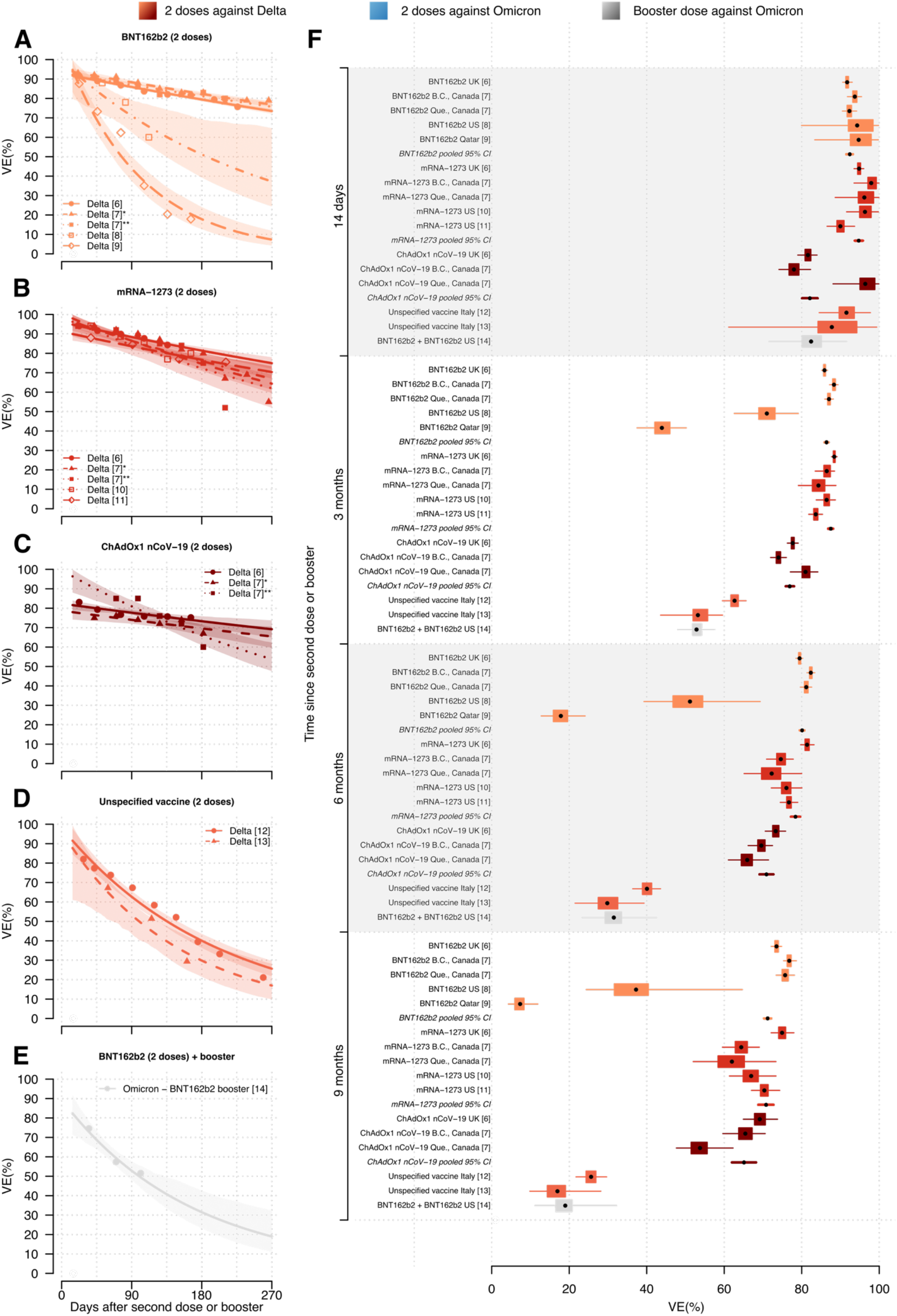
Effectiveness over time of two-dose and booster vaccination against any Delta and Omicron laboratory-confirmed symptomatic infections. **A-D)** Estimated vaccine effectiveness (VE) over time against any laboratory confirmed SARS-CoV-2 infection with Delta after administration of two doses of BNT162b2 (A), mRNA-1273 (B), ChAdOx1 nCoV-19 (C), and unspecified COVID-19 vaccine (D). Lines: mean estimates; shaded areas: 95% CIs; points: original VE estimates from published articles [6-13] (placed at the midpoint of the time interval for which the estimate was obtained); *data from British Columbia; **data from Quebec. **E)** Estimated VE over time against any laboratory confirmed SARS-CoV-2 infection with Omicron after the administration of a booster dose of BNT162b2 following two doses of BNT162b2. Lines: mean estimates; shaded areas: 95% CI; points: original VE estimates from published article [14] (placed at the midpoint of the time interval for which the estimate was obtained). **F)** Comparison of VE against any laboratory confirmed SARS-CoV-2 infection with Delta and Omicron across different vaccine products at 14 days, and at 3, 6, and 9 months from the administration of second vaccine dose or booster dose. Points: mean estimates; boxes: interquartile ranges; whiskers: 95% CIs.

**Figure 2.**
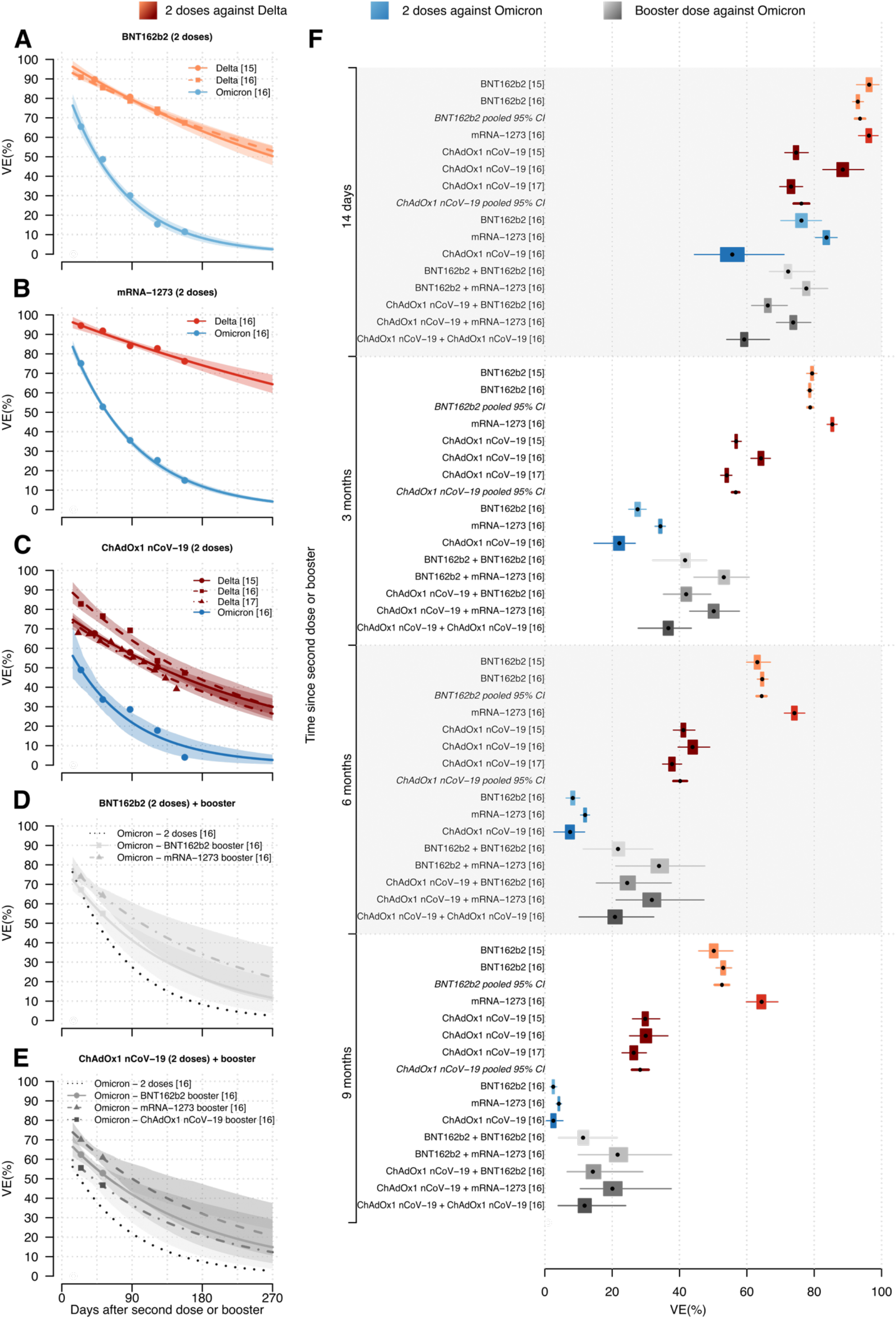
Effectiveness over time of two-dose and booster vaccination against Delta and Omicron symptomatic infections. **A-C)** Estimated vaccine effectiveness (VE) over time against symptomatic Delta and Omicron SARS-CoV-2 infection after the administration of two doses of BNT162b2 (A), mRNA-1273 (B), and ChAdOx1 nCoV-19 (C). Lines: mean estimates; shaded areas: 95% CIs; points: original VE estimates from published articles [15-17] (placed at the midpoint of the time interval for which the estimate was obtained). **D-E)** Estimated VE over time against symptomatic infection with Omicron after the administration of a booster dose following two doses of BNT162b2 (D) and of ChAdOx1 nCoV-19 (E). Lines: mean estimates; shaded areas: 95% CI; dashed black lines: mean VE after two doses of vaccine; points: original VE estimates from published article [16] (placed at the midpoint of the time interval for which the estimate was obtained). **F)** Comparison of VE against symptomatic SARS-CoV-2 infection with Delta and Omicron across different vaccine products at 14 days, and at 3, 6, and 9 months from the administration of second vaccine dose or booster dose. Points: mean estimates; boxes: interquartile ranges; whiskers: 95% CIs.

### VE against laboratory confirmed infection

Pooled estimates of VE 14 days after administration of the second vaccine dose BNT162b2 and mRNA-1273 against any SARS-CoV-2 laboratory confirmed infection (symptomatic or asymptomatic) with Delta are 92.4% (95%CI 91.5%-93.2%) and 94.7% (95%CI 93.7%-95.8%), respectively (see Figure 1, Table 1, and Figure S2). While a similar initial VE was found for ChAdOx1 nCoV-19 in Quebec (96.4%; 95%CI 88.1%-99.9%), estimates associated with studies conducted in the UK and in British Columbia suggest a lower VE for ChAdOx1 nCoV-19 compared to BNT162b2 and mRNA-1273 both soon after the administration of the second dose (mean estimates: 78.0%-81.6% vs 91.7%-93.7% and 94.8%-97.9%) and at 3 months from vaccination (mean estimates: 74%-77.7% vs 85.9%-88.3% and 86.5%-88.4%). However, no marked differences in the VE against Delta infection of mRNA-1273 and of ChAdOx1 nCoV-19 were found at 9 months from administration of the second dose (pooled estimates: 70.8% 95%CI: 68.9%-72.7% vs 65.1% 95%CI: 62.0%-68.2%). A marked uncertainty surrounds the temporal decline of protection provided by BNT162b2 against Delta infection, with mean estimates of VE at 9 months from administration of the second dose ranging from 7.3% to 76.8% (95%CI of pooled estimates: 70.3%-72.1%). Specifically, results based on VE estimates reported by studies conducted in US and Qatar suggest a significantly faster decline of VE over time associated to BNT162b2 (mean half-lives: 200 and 82 days in the two countries respectively; see Table 1). A similar waning of protection was found from data provided by the two Italian studies, where subjects vaccinated with one dose from less than 14 days were considered as a proxy for unvaccinated individuals [12,13]. In this case, the protection of two doses of unspecified vaccines against Delta infection was found to decrease from a mean VE ranging from 87.8% to 91.6% at 14 days from administration, to 29.8%-40.1% and 17.0%-25.6% at 6 and 9 months from administration, respectively. These estimates are in line with the VE associated with three doses of BNT162b2 against Omicron infection, which we estimated to be 82.4% (95%CI 71.5%-91.6%), 31.5% (95%CI 23.4%-42.5%) and 19.0% (95%CI 11.2%-32.2%) at 14 days, at 6 and 9 months from last dose administration, respectively.

### VE against symptomatic infection

Pooled estimates of VE 14 days after administration of the second vaccine dose BNT162b2 and ChAdOx1 nCoV-19 against symptomatic infection with Delta are 93.6% (95%CI 92.1%-95.1%) and 76.2% (95%CI 73.9%-78.5%; see Figure 2, Table 2, and Figure S2). The corresponding estimate for mRNA-1273 is 96.2% (95%CI 93.1%-99.0%). VE 14 days after administration of the second vaccine dose against symptomatic infection with Omicron was found to be 76.3% (95%CI 70.6%-82.4%), 56.1% (95%CI 44.4%-71.1%), and 83.6% (95%CI 80.4%-86.8%) with two doses of BNT162b2, ChAdOx1 nCoV-19, and mRNA-1273 respectively. At 6 months from the administration of the second dose, pooled estimates of VE against Delta variant for BNT162b2 and ChAdOx1 nCoV-19 declined to 64.4% (95%CI 62.9%-65.8%) and 40.2% (95%CI 38.2%-42.2%), respectively. The corresponding estimate for mRNA-1273 is 74.1% (95%CI 71.1%-77.2%). Our estimates of VE against symptomatic infection with Omicron at 6 months from the administration of two doses of BNT162b2, ChAdOx1 nCoV-19, and mRNA-1273 are 8.3% (95%CI 6.2%-10.4%), 7.5% (95%CI 2.7%-11.8%), and 12.0% (95%CI 10.6%-13.3%) respectively. The faster waning of protection provided by two doses against Omicron compared to Delta is evident when considering the half-life of vaccine-induced immunity against symptomatic infection, ranging on average from 178 to 456 days for Delta, and from 66 to 73 days for Omicron (Table 2). The estimated mean VE of two doses in preventing symptomatic infection with Omicron at 9 months from vaccination was less than 5% for any considered vaccine product.

Our estimates show that the booster dose has the potential of restoring VE at levels comparable with those acquired soon after the administration of the second dose (Figure 2 and Table 2). Despite we found that any booster dose increases the mean duration of the vaccine-induced protection compared with the administration of two doses only, our results suggest that at 9 months after administration of the last dose the mean VE of the booster dose against symptomatic infection with Omicron ranges between 11.7% and 22.2%. The estimated mean half-life of booster VE against symptomatic infection with Omicron ranges from 107 to 149 days (see Table 2).

## Discussion

We combined published evidence on the effectiveness of different vaccine products in preventing SARS-CoV-2 infection and COVID-19 symptoms [6-17] to estimate the duration of vaccine-induced protection against different variants. Results were used to quantify the level of vaccine-induced protection provided at any time from last dose administration, improving the comparability across different vaccines and number of doses over relatively longer time horizons.

The performed analysis highlighted that the effectiveness of vaccination with 2 doses of BNT162b2, mRNA-1273, and ChAdOx1 nCoV-19 against any laboratory confirmed infection with Delta might have been lower than 70% at 9 months from last dose administration, and that the protection provided by a booster dose against Omicron infection rapidly wanes over time (31.5% mean VE at 6 months from administration). We found that the emergence of the Omicron variant reduced the initial effectiveness acquired from the primary vaccination course against symptomatic infection as well (from 76.2%-93.6% to 56.1%-83.6%, depending on the product considered), while increasing the pace of waning of protection. Our mean estimates indicated that at 6 months from the second dose, any considered vaccine has an effectiveness of less than 13% against Omicron symptomatic infection. The administration of a booster dose was found to restore VE at levels comparable to those acquired after the second dose and to slow down the rate of waning. Compared to two doses, booster vaccination increased vaccine protection against symptomatic disease by 33.3%-56.3% at 3 months from the administration of the last dose. This result is in line with the 55% increase of VE against Omicron resulting from booster administration estimated by analyzing secondary attack rates in households [18]. However, our projections suggest that at 9 months from booster administration the mean VE against symptomatic infection with Omicron could be less than 25%.

These results should be interpreted by considering the following limitations. Our analysis is based on a small set of articles representing published studies up to June 21, 2022, providing estimates of the VE at different times from vaccine administration. In particular, VE against Omicron infection and symptomatic disease were inferred from two studies only [14,16]. Due to the limited duration of the follow-up period associated with the effectiveness of booster doses against Omicron, only two time points were used to inform our model estimates of VE against Omicron symptomatic infection. Additionally, original VE estimates against Omicron symptomatic infection [16] refer to a period in which the dominantly circulating sub-lineage was BA.1. VE against any laboratory infection with Omicron [14] were originally assessed in a period that encompasses the emergence of BA.2 sub-lineage as well. A recent study [19] showed that the waning of vaccine immunity after the booster dose was similar between Omicron sub-lineages BA.1 and BA.2. However, uncertainty remains on the effect of boosters against more recent sub-lineages. More in general, estimates of VE against laboratory confirmed infection should be also cautiously interpreted as they may not reflect immunity provided against any SARS-CoV-2 infection. In fact, preferential testing on symptomatic individuals may have biased the original assessment of the vaccine-induced protection against the infection. Different study designs were associated with the original estimates used for model calibration. This includes the heterogeneous definition of the reference group (unvaccinated vs vaccinated from less than 14 days) and the type of study (retrospective cohort vs case-control studies). This might have affected the observed differences among the original estimates of the vaccine effectiveness (e.g., a more rapid waning of immunity in the study with the reference group made of recently vaccinated individuals). Finally, current evidence suggests that natural and hybrid immunity (one dose of vaccine followed by recovery from infection, or recovery from infection followed by one dose of vaccine) might be more durable than vaccine-induced immunity [20]. Due to lack of data, this study did not investigate the possible differences in waning of vaccine protection for individuals who have never experienced a SARS-CoV-2 infection compared to previously infected individuals. Given the marked increase of breakthrough infections after the emergence of the Omicron variant [3-5], comparing duration of vaccine-induced and natural immunity remains a relevant open issue.

The estimates provided in this study can be instrumental to evaluate the expected susceptibility profile of different populations, while encouraging the discussion on appropriate targets and timing for future vaccination programs. In principle, our results highlighted that a marked immune escape is associated with Omicron infection and symptomatic disease both after two doses and after booster administration. However, booster doses were found to restore the vaccine protection against symptomatic disease to levels comparable to those estimated soon after administration of the second dose, while reducing the pace of its waning. Given the persisting circulation of SARS-CoV-2 observed in most high-income countries and the potential emergence of new Omicron sub-lineages or other SARS-CoV-2 variants in the future, additional vaccination efforts may be required to avoid a resurgence of COVID-19 symptomatic patients.

## Data Availability

All data produced in the present work are contained in the manuscript

## Acknowledgements

P.P. and V.M. acknowledge funding from EU grant 694160 VERDI. S.M., G.G., M.M. acknowledge funding from EU grant 874850 MOOD. The funders had no role in study design, data collection and analysis, decision to publish, or preparation of the manuscript.

## Authors’ contributions

P.P., M.A., and S.M. conceived the study. F.M. and P.P. wrote the first draft of the manuscript. F.M. wrote the code and performed the analyses. P.P., M.A., and S.M. supervised the study. F.M., P.P., M.M., G.G., V.M., A.Z., Vd.A., F.T., M.A., and S.M. interpreted results. All authors read, reviewed, and approved the final version and the submission of the manuscript. The corresponding author had final responsibility for the decision to submit for publication

## Supplementary material

**Table S1.**
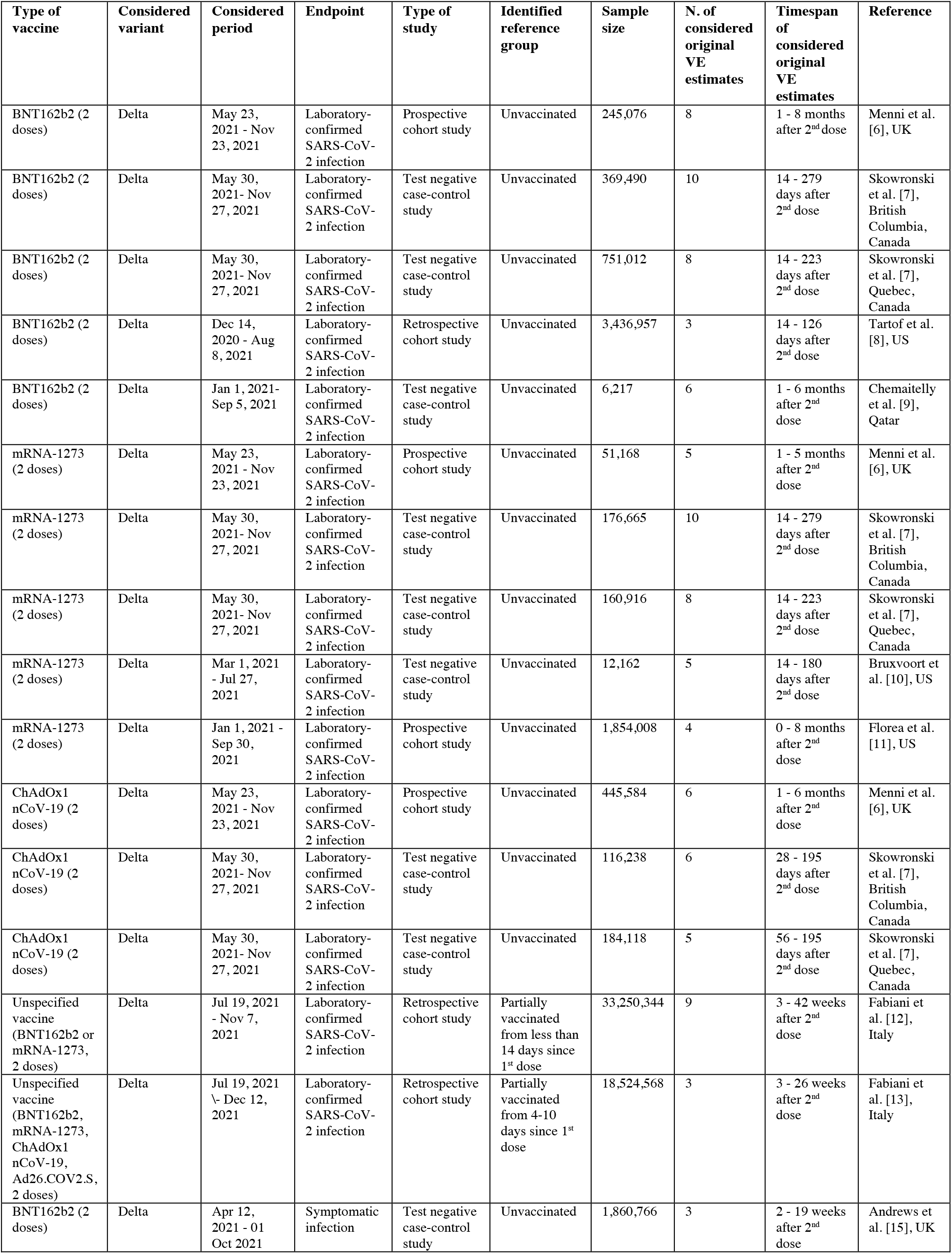
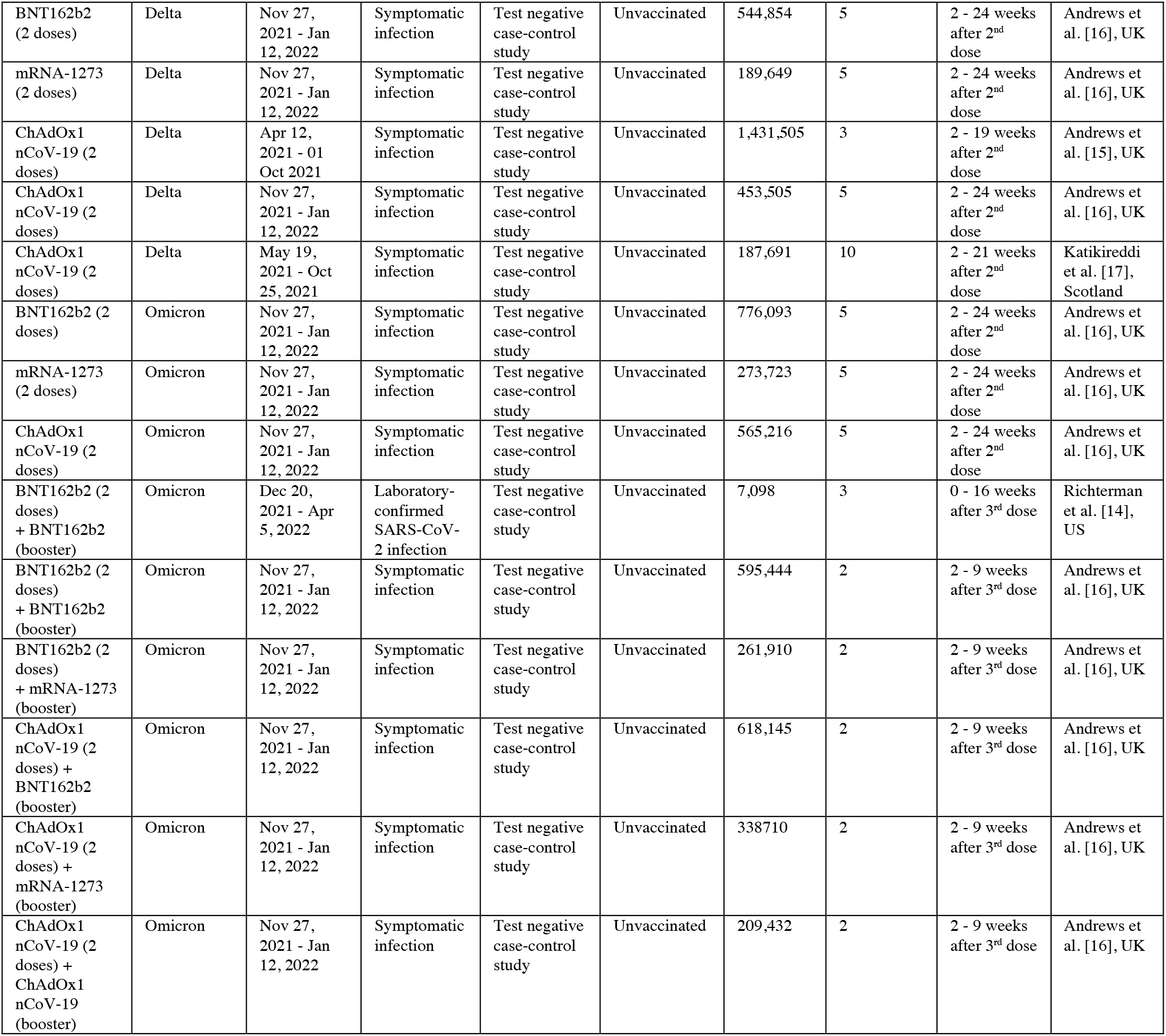
Sample size, study period, type of study, lineage, vaccine product, number of doses, and outcome associated with the analyzed time series of VE.

**Figure S1.**
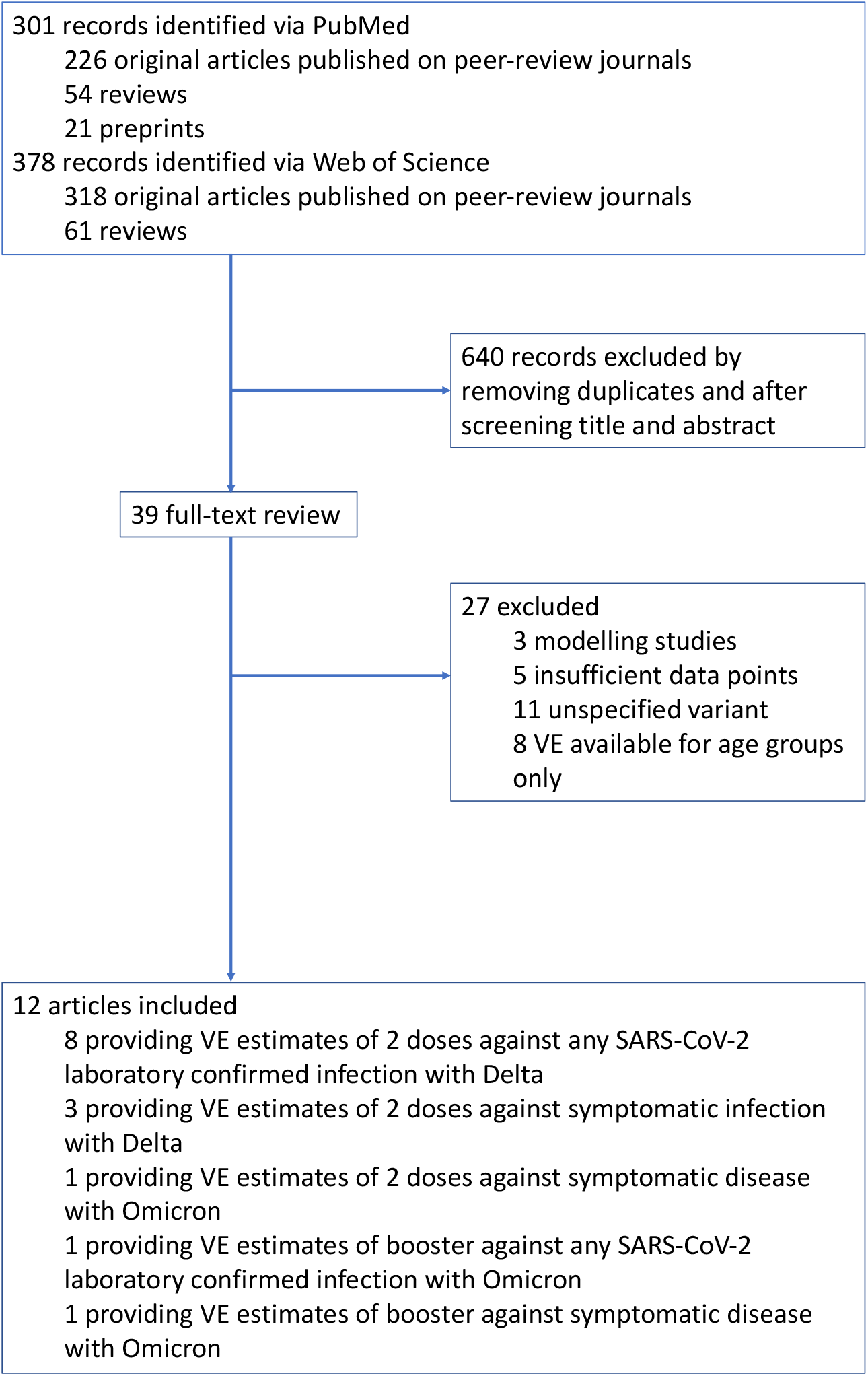
Study selection. Flowchart of the selection of studies considered for the performed analysis.

**Figure S2.**
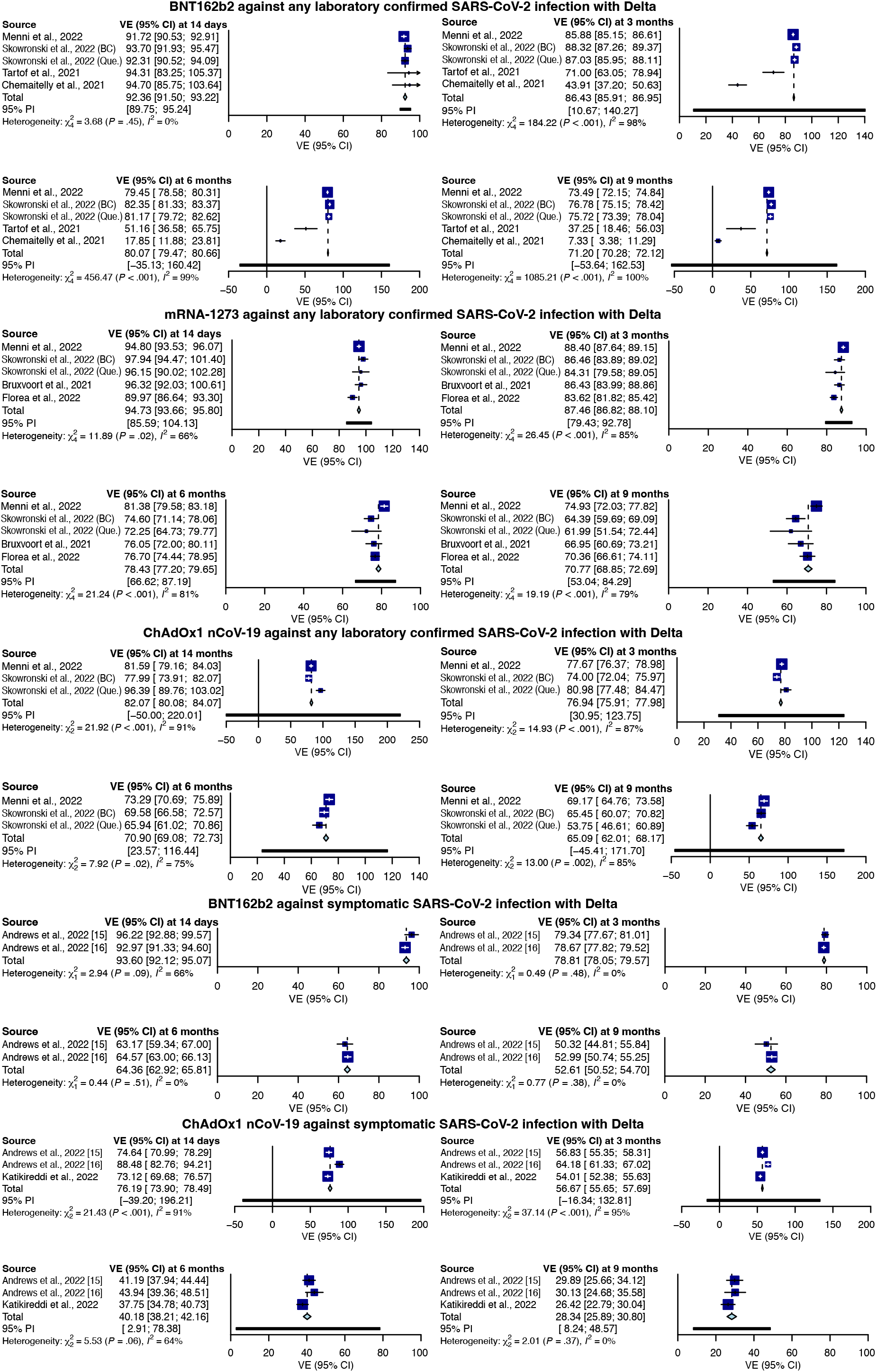
Pooled estimates obtained with the meta package (R statistical software version 4.1.2).

